# Validation of a novel scale for outcome measurement in Functional Neurological Disorder: The Functional Neurological Disorders Rating Scale (FuNDRS)

**DOI:** 10.1101/2025.10.23.25338345

**Authors:** Lucy Miller, Gurjot Brar, Gabriella Caceres Gamundi, Lisette Guy, Simon Harrison, Nick Medford, Paul Shotbolt

## Abstract

Functional Neurological Disorder (FND) is a common and disabling condition lacking an outcome measurement tool to guide efficacies of interventions in clinical trials. The Functional Neurological Disorders Rating Scale (FuNDRS) is a 25-item clinician-rated scale that generates scores across all symptom domains in FND and is designed to detect true symptom progression with sensitivity to change. This study demonstrates the initial validity of the FuNDRS using convergent, concurrent, discriminant, predictive and known-groups validity. Additionally, factor analysis, internal consistency, sensitivity to change and inter-rater reliability analyses are reported to further accrue evidence for validity. Collection of data in the day hospital arm (n=59) included FuNDRS, HoNOS, CGI-I, BDI, BAI, WSAS, PHQ-9 and GAD-7. Collection of data in the inpatient arm (n = 39) included FuNDRS, CGI S and CGI-I. Data analysis was performed using Pearson’s correlations, t-tests, quadratic weighted kappa and intra-class coefficient, and a principal components analysis. The FuNDRS has good internal consistency (α = 0.81) and showed overall strong positive correlation with the HoNOS and CGI (r > 0.6). Most items assessed for concurrent and discriminant validity displayed moderate to strong correlation. Patients scored lower at discharge, reflecting a statistically significant average difference of -12.41 (SE = 0.83) BCa 95% CI [10.77,14.05], resulting in a very large effect size of d = 8.19. The MDC95 was 2.3. Inter-rater reliability analysis was conducted on a subsample of inpatients found statistically significant agreement between two raters. Substantial agreement (Weighted Kappa > 0.8) occurred for change in scores on the FuNDRS and CGI scales. These findings support the initial validity of the FuNDRS as an outcome measurement tool for FND.

## Introduction

Functional Neurological Disorder (FND) is characterised by cognitive, motor and/or sensory symptoms that do not correlate with clinical features of neurological/medical disease. There is no known organic cause identified by routine investigations. The disorder is associated with significant distress and functional impairment. Common symptoms include sensory and motor phenomena, perceived cognitive issues, pain and non-epileptic seizures (American Psychiatric Association, 2013; Espay et al., 2018).

Due to the marked heterogeneity and variability in FND symptoms, it can prove challenging to define and measure outcomes (Nicholson et al., 2020). A prominent feature is the disparity between objective measures and subjective experiences (Pareés et al., 2012), also seen in mental disorders (Pies, 2007) and pain (Wideman et al., 2019). However, this disparity in FND diminishes in more severely disabled patients, such as those admitted to specialised inpatient units (Nicholson et al., 2020). Providing further support to objectivity in FND diagnosis, it is no longer necessary for clinicians to demonstrate presence of psychological stressors or absence of malingering in the patient (American Psychiatric Association, 2013). This freeing clause allows for symptoms to be taken at face-value and rated objectively by clinicians.

In addition to the core features of FND, several other physical and psychological symptoms are often reported and correlate with a reduced quality of life and greater disability (LaFrance et al., 2009; Jones et al., 2016; Gelauff et al., 2018); in some cases, more so than the presenting complaint (Věchetová et al., 2018). These symptoms are varied and include pain, fatigue, bladder and bowel dysfunction, and psychological symptoms, such as depression and anxiety (Stone et al., 2010a; Kranick et al., 2011). Correspondingly, the elevated symptom burden carries greater disability, role impairment and ultimately increased healthcare use and expenditure (Barsky et al., 2005; Harris et al., 2009).

Initial management involves appropriate diagnostic delivery and due deference to possible functional overlay symptoms. Treatment modalities can generally be divided into the psychological and physical. Additionally, co-morbid depression, anxiety and pain can be treated by conventional pharmacological methods. Psychological interventions have shown promising results for non-epileptic seizures (Goldstein et al., 2010; LaFrance et al., 2014) and motor symptoms (Moene et al., 2003). Physical interventions have also recently developed an evidence base (Jordbru et al., 2014; Nielsen et al., 2015; 2017), and it has now been suggested that integrating psychotherapeutic modalities in a multidisciplinary context offers the most success (Espay et al., 2018). More treatment modalities are being explored and require more robust findings before their application can be recommended (Espay et al., 2018; Gilmour et al., 2020). Despite the ground-breaking work from various disciplines done in this area, these studies lack a common outcome measurement tool for adequate comparison.

Consensus on outcome measures is still lacking despite the recent progression of FND in research and FND symptoms being among the most common presentations to neurology clinics after headaches (Carson et al., 2000; Stone et al., 2010b). There is no single measure identified for use across the range of FND symptoms in adults (Pick et al., 2020). Previous tools which measure the outcome of specific deficits only are hampered by the nature of FND patients to develop new, additional functional symptoms. Indeed, as Gelauff & Stone (2016) have described, having a functional symptom increases the risk of developing other functional symptoms. There are many challenges to designing such a tool for FND which presents with a wide range of symptom variability and can mimic several neurological disorders (Nicholson et al., 2020). The FND Core Outcome Measurement group (FND-COM, 2017) represents an initiative in the wider Core Outcome Measures in Effectiveness Trials group (COMET) to standardise outcome measurement across trials.

The Functional Neurological Disorders Rating Scale (FuNDRS) is a novel clinician-rated single outcome measurement tool that has been developed with the aim of standardising outcome measurement in FND by our FND working group at the Institute of Psychiatry, Psychology and Neuroscience, King’s College London. The FuNDRS generates scores across all symptom domains in FND and is designed to detect true symptom progression with sensitivity to change. It consists of the five following domains: mental status, impact of illness, motor function, sensory symptoms, and other associated symptoms Establishing a gold-standard measurement tool for outcomes in FND will have several benefits: improving consistency across research studies assessing interventions, providing comparable therapeutic indices of interventions, fostering support for clinical decision; and finally, providing information to healthcare providers regarding costs to justify or discontinue services and interventions (Slade et al., 2006).

Outcome measurement tools require validity in target populations to be credible (Boers et al., 1998; de Vet et al., 2011). Validity describes whether an instrument measures what it claims to measure (Cronbach, 1971). In that respect, the FuNDRS has potential to be a valid tool as it measures FND outcomes across multiple symptom domains in target populations. Construct and concurrent validity describe the amount of agreement between established tools and nascent ones. When evaluating a new instrument, high concurrent validity with a well-validated criterion is desirable as it indicates a valid test instrument (Miller et al., 2011). Multidimensional scales measuring outcomes for Parkinson’s Disease (Movement Disorder Society revision of the Unified Parkinson’s Disease Rating Scale (MDS-UPDRS) (Goetz et al., 2008), Huntington’s Disease (Unified Huntington’s Disease Rating Scale (HDRS) (Kieburtz et al., 2001) and obsessive-compulsive disorder (Yale-Brown Obsessive Compulsive Scale (YBOCS) (G oipioodman et al., 1989) have been previously developed and validated, leading to their widespread use.

In this paper, we perform validation of the FuNDRS against two established outcome measurement tools. The two most appropriate tools that were identified were the Health of the Nation Outcome Scales (HoNOS) (Wing et al., 1998) and the Clinical Global Impression Scale (CGI) (Guy, 1976; Busner & Targum, 2007).

### HoNOS

The HoNOS is an extensively administered 12-item clinician-rated outcome measure for adults in the UK attending mental health services. It has been validated previously for assessment of psychiatric symptoms, behaviour, and social functioning (James et al., 2018). The HoNOS has been demonstrated to capture sensitivity to change in various symptom domains of FND, with HoNOS scores also predicting the level at which patients’ self-rate their improvement (Wing et al. 1998; Demartini et al., 2014).

### CGI

The CGI is a well-known and validated brief clinician-rated tool used typically in research assessing efficacy of treatments in psychiatric disorders (Leon et al., 1993; Zaider et al., 2003; Hale et al., 2010). The CGI measures severity of illness, therapeutic response, and global improvement. The CGI has aided validation studies by providing an external criterion to assess validity of other novel outcome measures in the past (Lindenmayer et al., 2003; Korner et al., 2007; Riedel et al., 2010;).

Thus, the FuNDRS will be validated against the HoNOS and CGI to determine if it is also a valid instrument of outcome measurement. The FuNDRS will also be validated through comparison with the following outcome scales: Beck Depression Inventory-II (BDI-II) (Beck et al., 1996); Beck Anxiety Inventory (BAI) (Beck et al., 1988); Work and Social Adjustment Scale (WSAS) (Mundt et al., 2002); Patient Health Questionnaire (PHQ-9) (Kroenke & Spitzer, 2002) and the Generalised Anxiety Disorder Scale (GAD-7) (Spitzer et al., 2006). Additionally, concurrent and discriminant validity, factor analysis, sensitivity to change and inter-rater reliability of the FuNDRS will be assessed to further accrue evidence for validity. By validating this novel scale, it is hoped that it will become widely used both for evaluation of FND treatment services and to demonstrate efficacy in clinical trials of new interventions.

## Method

### Sample

There were two conceivable patient arms in this study determined by a convenience sample of patients admitted to either arm over a certain period. The first arm were all consecutive patients diagnosed with FND who are enrolled on the Functional Integrated Neurological Disorders (FiND) outpatient day programme at the Bethlem Royal Hospital (South London & Maudsley NHS Foundation Trust, London, UK) (from here on known as the ‘FiND arm’), from May 2018 to March 2020 and then from May 2022 to July 2022. The reason for the hiatus was COVID-19 pandemic-related. The second arm consisted of all consecutive patients diagnosed and admitted with FND at the Lishman Unit, Bethlem Royal Hospital (South London & Maudsley NHS Foundation Trust, London, UK) (from here on known as ‘Lishman Unit arm’) from May 2017 to March 2020. There were no new FND admissions to the ward between March and September 2020 for reasons related to the COVID-19 pandemic.

The Lishman Unit is a 15-bedded specialist inpatient neuropsychiatry unit located at the Bethlem Royal Hospital. At any given time around half the beds are allocated to patients with severe FND, and the ward is one of only a handful of units in the UK that admits such patients for physical and psychological treatment and rehabilitation. A typical admission lasts 3-6 months. During this period patients receive regular sessions of physiotherapy, occupational therapy, clinical psychology, and where indicated, speech and language therapy, in additional to medical and nursing care. Patients referred to the unit generally have severe FND, often having a combination of longstanding symptoms (e.g., motor symptoms, cognitive symptoms, and non-epileptic seizures) and significant disability. The basic multidisciplinary therapy approach is adapted for each individual case through team discussion and joint working. A similar approach is undertaken in the FiND arm, where a structured day hospital programme is provided. Cohorts of 3-6 FND patients are given multimodal therapy, primarily by occupational therapy and psychology for two days a week over a period of 8 weeks.

Across both arms, data was collected from a total sample of 98 patients (59 from FiND and 39 from the Lishman Unit). Although no general criteria for sample size exists for validation of instruments (Streiner et al., 2015), a previous assumption recommends the number of participants to exceed the number of items by a factor of three (Barrett & Kline, 1981). As the FuNDRS has 25 items, a sample of 98 is more than adequate.

### Procedure

Clinician-rated and patient-reported measures were taken at two intervals: T1 – at patient admission to treatment programme (FiND arm) or psychiatric unit (Lishman Unit arm); and T2 – at patient discharge. Collection of data in the FiND arm included FuNDRS, HoNOS, CGI-I, BDI, BAI, WSAS, PHQ-9 and GAD-7. Collection of data in the Lishman arm included FuNDRS, CGI-S and CGI-I.

The investigator PS independently rated FuNDRS scores at T1 and T2 using retrospective case notes for a sample of 18 patients from the Lishman arm.

The rating scales and outcome measures used in this study are administered within 48 hours of admission, the purpose of these being explained with patients given a free choice as to whether they complete these measures. They serve the purpose of enabling the ward team to assess the initial symptom burden, and to track progress during the admission. Rating scale completion is repeated at various time points during the admission, and again at discharge. In this study, data from admission and discharge, but not from intermediate time points, is presented.

### Ethics

All data was collected as part of the normal working conditions of the FiND programme and the Lishman Unit at the South London and Maudsley NHS Foundation Trust by trained mental health professionals. The data presented in this study were obtained from anonymised clinical records originally collected for service evaluation and quality improvement purposes.

No identifiable patient information was used, and the project did not alter patient care. Thus, no informed consent was required or obtained.

## Measures

### Functional Neurological Disorders Rating Scale (FuNDRS)

The FuNDRS is a clinician-rated outcome measurement tool consisting of twenty-five items across five domains, reflecting the symptom clusters of FND: Mental State’ (five items), ‘Impact of Illness’ (four items), ‘Motor Function’ (six items), ‘Sensory Symptoms’ (six symptoms) and ‘Other Associated Symptoms’ (such as bladder dysfunction) (four items).

Each item is scored on a 5-point severity scale ranging from 0 to 4: 0 = no symptoms/problems; 1 = minor; 2 = mild;3 = moderate; 4 = severe. The clinician rates the score for each item based on symptoms over the last two weeks, with a greater score reflecting high symptom severity. The maximum score is 100.

### Health of Nations Outcome Scales (HoNOS)

The HoNOS is an extensively administered 12-item clinician-rated outcome measure for adults in the UK attending mental health services. It has been validated previously for assessment of psychiatric symptoms, behaviour, and social functioning (James et al., 2018). Each item is scored on a 5-point scale ranging from 0 to 4 to represent maximum severity over the rating period over the previous 2 weeks: 0 = no problem; 1 = minor problem requiring no action; 2 = mild problem but definitely present; 3 = moderately severe problem; 4 = severe to very severe problem. A higher score therefore indicates greater symptom severity, with the maximum score being 48.

### Clinical Global Impression (CGI)

This is a 3-item clinician-rated instrument that measures illness severity, global improvement/ change, and treatment efficacy. Item 1 (CGI-S) measures the Severity of Illness and is rated on a 7-point scale ranging from 1 (normal) to 7 (among the most extremely ill patients); item 2 (CGI-I) measures the patients global improvement after a course of treatment and is rated on a 7-point scale ranging from 1 (very much improved) to 7 (very much worse); item 3 refers to the efficacy index of therapeutic response and is rated on the basis of drug effect only and is rated from 1 (marked improvement and no side effects) to 4 (unchanged/worse and side effects outweigh the therapeutic effects). A higher score therefore indicates greater severity/ worse outcomes. Item 3 relating to efficacy of therapeutic response was not used in this study as it relates to drug effect only and both arms are provided with comprehensive multi-disciplinary management.

### Beck Depression Inventory-II (BDI-II)

The BDI-II is a validated, extensively used 21-item self-reporting questionnaire used in evaluating the severity of depression. Each item is scored on a 4-point severity scale ranging from 0 (symptom absent) to 3 (severe symptoms) based on the patient’s symptoms over the previous two weeks. A higher score therefore indicates greater symptom severity/depression level, with the maximum score being 63: 1 - 10 (normal); 11 - 16 (mild mood disturbance); 17 - 20 (borderline clinical depression); 21 - 30 (moderate depression); 31 - 40 (severe depression); over 40 (extreme depression).

### Beck Anxiety Inventory (BAI)

The BAI is a validated, extensively used 21-item self-reporting questionnaire used in evaluating the severity of anxiety symptoms in the past week. Each item is rated on a 4-point severity scale ranging from 0 to 3: Not at All = 0; Mildly = 1; Moderately = 2, and Severely = 3. Therefore, higher scores indicate greater symptom severity/anxiety levels, with the maximum total score being 63: 0 - 7 (minimal anxiety); 8 - 15 (mild anxiety); 16 - 25 (moderate anxiety); 26 - 63 (severe anxiety).

### Work and Social Adjustment Scale (WSAS)

The WSAS is a 5-item self-reporting questionnaire evaluating the impact of mental health difficulties and subsequent impairment in functioning. Each item corresponds to a different impairment dimension: work; home management; social leisure activities; private leisure activities; relationships with others. Each item is scored on an 8-point severity rating scale ranging from 0 (not at all) to 8 (very severely). Therefore, a higher score indicates worse adjustment, with the maximum total score being 40: below 10 (subclinical); 10 - 20 (significant impairment with less severe clinical symptomatology); above 20 (moderate to severe impairment).

### Patient Health Questionnaire (PHQ-9)

The PHQ-9 is a 9-item self-administered questionnaire for depressive disorder. Diagnosis can be made if 5 of 9 depressive symptom criteria are present for at least “more than half the days”. The score ranges from 0 to 27 and each item can be scored from 0 (“not at all”) to 3 (“nearly every day”): 5-9 (minimal symptoms), 10-14 (Minor depression, dysthymia, or major depression, mild), 15-19 (Major depression, moderately severe) and 20 or greater (Major Depression, severe).

### Generalised Anxiety Disorder Scale (GAD-7)

The GAD-7 is a shorter 7-item self-administered version of the 13-item questionnaire initially developed based on the DSM-IV diagnostic criteria of GAD (Spitzer, 2006). Although initially developed for GAD, it has been increasingly used for anxiety in general (Beard and Bjorgvinsson, 2014). The score ranges from 0 to 21, although a score of 8 or higher is a reasonable cut-off for further evaluation of a possible anxiety disorder: 0-4 (Minimal), 5-9 (Mild), 10-14 (Moderate), 15 or greater (Severe).

### Statistical Analyses

All analyses were conducted using IBM SPSS Statistics 28 (2021). The purposeful (convenient) nature of sample selection precluded meaningful estimates of scale-skewness and floor and ceiling effects to be computed (Hays et al., 1993).

### Overview of Validity Assessment

To ensure that the FuNDRS is a psychometrically sound tool, multiple forms of validity were assessed. *Construct validity* refers to the extent to which the scale measures the theoretical construct it was designed to assess. This includes *convergent validity* (the degree of correlation with other instruments measuring similar constructs), *discriminant validity* (low correlation with unrelated constructs), and *known-groups validity* (ability to distinguish between groups known to differ on the construct). *Criterion validity* was explored through *concurrent validity* (association with established “gold standard” measures completed simultaneously), while *sensitivity to change* evaluates the ability of the scale to detect meaningful clinical improvement over time.

### Internal Consistency

To be established by Cronbach’s alpha which measures internal consistency and is a prerequisite to factor analysis. Internal consistency describes the extent to which all the items in a test measure the same concept or construct and hence it is connected to the inter relatedness of the items within the test. Internal consistency should be determined before a test can be employed for research or examination purposes to ensure validity. (Cronbach 1971; Tavakol & Dennick, 2011).

### Convergent Validity

Convergent validity is demonstrated by comparing measures of instruments measuring similar but different constructs. In that respect, the HoNOS and CGI, although measures of outcome, do not specifically measure outcomes related to FND.

Pearson’s correlation analysis allowed us to explore the relationship between scores on the FuNDRS with the HoNOS and the CGI. Each was performed for admission (T1), discharge (T2) and change in scores (T1-T2). Values of r = 0.5 and above signify a strong positive relationship, whereas those below r = 0.3 reflect a lack of convergent validity and are thus considered weak. Values of r = 0.5 and above are expected for T1, T2 and T1-T2 for both patient arms. Statistical significance is to be defined at the 5% threshold level.

### Discriminant Validity

Discriminant validity is useful to demonstrate the specificity of the instrument in question. Here it refers to the extent and degree to which two constructs differ from one another.

i. Depression was to be discriminated by comparing the ‘depressed mood’ item on FuNDRS with the total score on the WSAS and BAI.
ii. Anxiety was to be discriminated by comparing the ‘anxiety item’ on the FuNDRS with the total score on the WSAS and BDI.
iii. Functional Impact was to be discriminated by comparing all individual scores of items in the ‘Impact of Illness’ domain with the BAI and BDI.
iv. Gait was to be discriminated by comparing the ‘gait/mobility’ item on the FuNDRS with the score on the BDI and BAI.

It was expected that all discriminant values describe low correlations with their respective diverging measures. Correlations were to be regarded weak if less than or equal to 0.3 (Cohen, 1988).

### Known-groups Validity

To exhibit known-groups validity, an instrument should be able to discriminate between distinct groups of participants (Kerlinger, 1973). Known-groups validity was explored by investigating the ability of the FuNDRS to discriminate between patients with differences in their symptom improvement. The thresholds for determination were: 1) percent changes of the patients’ scores from T1 to T2, 2) percent changes grouped based on improvement status on the CGI-I, 3) averages of percent changes calculated for each improvement status category, 4) averages rounded to their nearest ten, and finally 5) cut-off thresholds created. Known-groups validity is quantified using independent samples t-tests at the 5% significance threshold.

**Table 1.** Known-Group Validity Thresholds and Score Reductions for FuNDRS scores based on CGI-I.

| <u>Threshold</u> | <u>Score Reduction</u> |
| --- | --- |
| Very much improved | $\geq 60\%$ |
| Much improved | 30% - 59% |
| Minimally improved | 1% - 29% |

### Factor Analysis

Factor Analysis is a statistical tool that can further assess the internal structure and construct validity of rating scales (Henson & Roberts, 2006; Kieffer, 1999). It was assumed that the factors would not correlate as FND and the FuNDRS are multidimensional in essence. Here we aimed to use principal axis factoring and orthogonal rotation on the FuNDRS items to demonstrate initial validity of the individual items to corresponding domains.

### Concurrent Validity

Concurrent validity can be demonstrated if a measure on the instrument in question correlates with those of a reference standard measurement tool, well known to accurately measure the same construct (Evans et al., 2002). For this, the ‘Depressed Mood’ item on the FuNDRS was compared with the total score of the BDI and PHQ-9 to demonstrate concurrent validity for depression. For anxiety, the ‘Anxiety’ item on the FuNDRS was compared with the total score on the BAI and GAD-7, and for functional impact, the ‘Impact of Illness’ domain (items 6-9) with the total score on the WSAS. Pearson’s correlational analysis will allow for the presence or absence of concurrent validity. Correlations are expected to be strong and thus ≥0.5 (Cohen, 1998) with 5% significant threshold are expected.

### Sensitivity to Change

The degree to which an instrument is capable in detecting change over time, as measured by against established gold standards, defines sensitivity to change (Ebesutani et al., 2012).

Paired-t tests allow for comparison of total scores of the FuNDRS, HoNOS and CGI-S at T1 and T2 to demonstrate responsivity to change.

The minimal detectable change at 95% confidence (MDC_95_) is to be calculated by the following formula: Standard Error of Measurement X 1.96 X √2 (Ries et al., 2009).

### Inter-rater Reliability

The investigator PS independently rated FuNDRS scores at T1 and T2 using retrospective case notes for a sample of 18 patients from the Lishman Unit arm. Cohen’s kappa (κ) was used to identify inter-rater reliability (IRR). It has the advantage of correcting for any “chance” agreement between the reviewers while also allowing for different types of disagreements to have different weights (Cohen, 1968). Additionally intra-class coefficient analysis with a two-way mixed model was conducted to further ascertain inter-rater reliability. The agreements among reviewers were categorised as follows (Landis & Koch, 1977): poor (0), slight (0.1-0.2), fair (0.21-0.4), moderate (0.41-0.6), substantial (0.61-0.8), or near perfect (0.81-0.99).

### Predictive Validity

If the FuNDRS demonstrates good sensitivity to change, a linear regression analysis can be used to predict FuNDRS scores at discharge using scores at admission.

## Results

The following section presents the psychometric analyses undertaken to examine the reliability and validity of the FuNDRS across inpatient and outpatient FND samples.

### Internal Consistency

In addition to adequate content and face validity, the FuNDRS has good internal consistency (α = 0.81) which describes the extent to which all the items in a test measure the same concept or construct and hence is connected to the inter-relatedness of the items within the test.

### Convergent Validity

Correlation analysis of the FiND arm with HoNOS scores, and the Lishman Unit arm with CGI-S scores at admission (T1) and discharge (T2) was conducted. Data for CGI-I were available from both arms (n = 98) and combined to correlate the change in scores on the FuNDRS with the CGI-I which yielded a Pearson product moment correlation coefficient of r = -0.618 (p<0.001), interpreted as strong correlation. Additionally, the FuNDRS displayed strong correlations (r exceeding 0.6) with the HoNOS at admission (T1), discharge (T2) and change (T1-T2) in the FiND arm (n=59). In the Lishman Unit arm (n=39), the FuNDRS displayed strong positive correlation with the CGI-S at discharge and change, and moderate positive correlation at admission. Thus, the FuNDRS has favourable convergent properties with the HoNOS, CGI-S and CGI-I. Further data including convergent data on domains and non-epileptic seizures (NES) is available in Table 2.

**Table 2.** Convergent Validity. Correlations at TI (admission), T2 (discharge) and change in score from T1 to T2. Combined n98; FiND n=59; Lishman n = 39.

|  | HoNOS<br>T1<br>(FiND) | HoNOS<br>T2<br>(FiND) | HoNOS<br>T1-T2<br>(FiND) | CGI-S T1<br>(Lishman) | CGI-S T2<br>(Lishman) | CGI-I T1-T2<br>(Lishman) | CGI-I<br>(Lishman) | CGI-I<br>(FiND) | CGI-I<br>(Combined) |
| --- | --- | --- | --- | --- | --- | --- | --- | --- | --- |
| FuNDRS T1 | 0.811** |  |  | 0.338* |  |  |  |  |  |
| FuNDRS T2 |  | 0.875** |  |  | 0.790** |  |  |  |  |
| FuNDRS T1-T2 |  |  | 0.689** |  |  | 0.790** | -0.644** | -0.595** | -0.618** |
| FuNDRS Domain 1 |  |  |  |  |  | 0.771** | -0.568** | -0.69 |  |
| FuNDRS Domain 2 |  |  |  |  |  | 0.814** | -0.705** | -0.261* |  |
| FuNDRS Domain 3 |  |  |  |  |  | 0.433* | -0.330* | 0.225 |  |
| FuNDRS Domain 4 |  |  |  |  |  | 0.559** | -0.443* | -0.214 |  |
| FuNDRS Domain 5 |  |  |  |  |  | 0.552** | -0.573** | -0.122 |  |
| NES T1 |  |  |  | 0.12 |  |  |  |  |  |
| NES T2 |  |  |  |  | 0.361* |  |  |  |  |
| NES T1-T2 |  |  |  |  |  | 0.328* | -0.416* | -0.169 |  |
| HoNOS T1-T2<br>(FiND) |  |  |  |  |  |  |  | -0.462** |  |

\* $p < 0.05$ \*\* $p < 0.001$
Note. FuNDRS = Functional Neurological Disorders Rating Scale; HoNOS = Health of Nation Outcomes Scale; CGI = Clinical Global Impression (Scale); CGI-S = Clinical Global Impression – Severity; CGI-I = Clinical Global Impression – Global Improvement; FiND = Functional Integrated Neurological Disorders (outpatient day programme); NES = Non-Epileptic Seizures. Pearson's correlation coefficient $r = 0.1-0.3$ (Weak), $0.3-0.5$ (Moderate) and $>0.5$ (Strong)

### Discriminant Validity

Data from the FiND arm were used to establish discriminant validity by means of Pearson’s correlation coefficient. Several items did not have divergent scales conducted on the same patients, limiting the number of items available to be assessed.

Many FuNDRS items have weak correlations (0.01-0.29) with their discriminating scales. There are some significant moderate correlations (0.39-0.59) and some significant strong correlations (0.61-0.7), particularly at discharge (T2) for Item 1 ‘Depression’ and BAI, and Item 2 ‘Anxiety’ with the WSAS and BDI. The latter likely resulting from features of depression and anxiety to be often concomitant.

**Table 3.** Discriminant Validity. Correlation between FuNDRS Items and criterion measures at T1 (admission), T2 (discharge) and change in score from T1 to T2. (FiND arm)

| FuNDRS Item | Criterion Measures | *r* |  |  |
| --- | --- | --- | --- | --- |
|  |  | T1 | T2 | Change |
| Item 1 – Depressed Mood | WSAS | 0.473** | 0.594** | 0.216 |
|  | BAI | 0.575** | 0.697** | 0.373** |
| Item 2 – Anxiety | WSAS | 0.390* | 0.612** | 0.421** |
|  | BDI | 0.579** | 0.682** | 0.414* |
| Items 6-9 (Impact Domain) | BAI | 0.287* | 0.623** | 0.035 |
|  | BDI | 0.174 | 0.569** | 0.240 |
| Item 15 – Gait/Mobility | BAI | -0.104 | 0.239 | 0.014 |
|  | BDI | -0.143 | 0.119 | 0.120 |

\* $p < 0.05$ \*\* $p < 0.001$
*Note.* FuNDRS = Functional Neurological Disorders Rating Scale; WSAS = Work and Social Adjustment Scale; BAI = Beck Anxiety Inventory; BDI = Beck Depression Inventory

### Known-Groups Validity

An independent-samples t-test indicated that scores were significantly higher for ‘very much improved’ patients (n=12, M= -24.42, SD = 6.97) than ‘much improved’ patients (n=33, M=16.64, SD = 6.08), a statistically significant difference of -7.78 (95% CI, -2.99 to -12.57), t(17.46) = 3.421, p = 0.003. Additionally, patients in the ‘much improved’ category had a significant reduction in scores compared to the ‘minimally improved’ patients (n= 51, M = - 7.33, SD = 4.17), with a statistically significant difference of -9.303 (95% CI, -6.88 to - 11.73), t(51.46) = 7.7, p < 0.001.

In total, only one patient in the FiND arm disimproved with a change in score of 2, reflecting a 4.88% increase. This analysis suggests the FuNDRS can detect differences in progress among patient groups.

### Factor Analysis

A sample size of 98 paired observations with a total of 206 observations, satisfied the criteria for at least five observations per variable (Comrey & Lee, 1992). In the Lishman arm, all 39 patients received admission and discharge scores whereas in the FiND arm, not all patients completed treatment or had a completed FuNDRS at discharge by the clinician (n = 10).

Thus, there is a discrepancy between admission and discharge numbers.

An initial principal components analysis was conducted on all 25 items with orthogonal rotation (Varimax). In multidimensionality, factors are not necessarily assumed to correlate, as in our case with FND presenting a multifaceted disorder. Initially 9 eigenvalues above 1 were obtained according to Kaiser’s criterion of 1 and in combination explained over 60% of the variance. This criterion can overestimate the number of factors to retain so a Scree plot is often consulted to interpret ideal factor retention, however it was ambiguous, and it would have been correct to assume that between four and nine factors were optimal.

A correlation matrix was consulted to identify any items with low correlation to other items, generally regarded as r<0.3. 7 of 25 items had r<0.3 correlation with other items and thus it was suggested to monitor these into the next stage of selection (Items 4, 5, 11, 12, 14, 17, 22). Kaiser-Meyer-Olkin (KMO) measure of sampling adequacy allows determination of appropriateness of running a principal components analysis. Initially, KMO was 0.716 and Bartlett’s test of sphericity was significant X (253) = 1267.5, p<0.001, indicating factor analysis is suitable. Further assessment of the Anti-image Correlation matrix using KMO measures for sampling adequacy revealed items 11, 22 and 24 had a KMO <0.5 which is deemed unacceptable for inclusion in factor analysis. These were subsequently removed and the KMO rose to 0.738 and factors suggested by eigenvalues >1 was 7. Further analysis with the interpretability criterion using the rotated component matrix, in addition to the eigenvalues and scree plot, determined a test of forced factor extraction be attempted at 5 factors. This yielded the most balanced interpretation of factor retention explaining 53.4% of the variance of the remaining 22 items.

Factor 1 consisted of ‘Mental State/Impact’, closely correlating depressive and anxiety symptoms with personal, social, and occupational functioning. Factor 2 consisted of ‘Physical Symptoms Impacting Mobility and Functioning’. Factor 3 consists of ‘Disorienting Symptoms’. Factor 4 consists of ‘Associated Symptoms’. Factor 5 relates to ‘Specific Motor Symptoms’ (see table 5 for individual items by factor).

Cross-loading occurred for eight items, in which case factor loading decisions were based on the larger value. However, for item 3 ‘Depersonalisation/Derealisation’ and item 9 ‘Problems with Relationships’, it made more conceptual sense to load these items on factors 4 and 1 respectively.

**Table 4.** Eigenvalues obtained from Principal Factor Analysis.

| **Component** | **Total** | **Variance (%)** | **Cumulative (%)** |
| --- | --- | --- | --- |
| 1 | 4.96 | 22.55 | 22.55 |
| 2 | 2.21 | 10.05 | 32.6 |
| 3 | 1.84 | 8.36 | 40.96 |
| 4 | 1.45 | 6.58 | 47.53 |
| 5 | 1.29 | 5.86 | 53.39 |

*Note. An eigenvalue >1 is considered meaningful according to Kaiser in interpretation of factor loading and retention.*

**Table 5.** Principal Factor Analysis of FuNDRS Items displaying factor items and their loadings (n=206)

|  |  | Rotated Factor Loadings |  |  |  |  |
| --- | --- | --- | --- | --- | --- | --- |
|  | FuNDRS Item | 1 | 2 | 3 | 4 | 5 |
|  | **1. Mental State/Impact** |  |  |  |  |  |
| 1 | Depressed Mood | 0.76 |  |  |  |  |
| 2 | Anxiety | 0.75 |  |  |  |  |
| 6 | Problems with ADL | 0.73 |  | [0.37] |  |  |
| 7 | Work | 0.65 |  | [0.37] |  |  |
| 8 | Hobbies/interests | 0.75 |  |  |  |  |
| 9 | Problems with relationships | 0.35 |  | [0.34] |  | [0.4] |
|  | **2. Physical Symptoms Impacting Mobility and Functioning** |  |  |  |  |  |
| 10 | Weakness |  | 0.78 |  |  |  |
| 15 | Gait/mobility | [0.43] | 0.45 | [0.32] |  |  |
| 16 | Loss of sensation/numbness |  | 0.75 |  |  |  |
| 25 | Daytime fatigue |  | 0.60 |  |  |  |
|  | **3. Disorienting Symptoms** |  |  |  |  |  |
| 4 | Cognitive |  |  | 0.49 | [0.31] |  |
| 18 | Sensitivity to light/sound |  |  | 0.71 |  | [0.32] |
| 19 | Dizziness |  |  | 0.70 |  |  |
|  | **4. Associated Symptoms** |  |  |  |  |  |
| 3 | Depersonalisation/Derealisation |  |  |  | 0.41 | [0.48] |
| 5 | Self-directed injury |  |  |  | 0.48 | [0.35] |
| 20 | Pain Symptoms |  | [0.45] |  | 0.57 |  |
| 21 | Analgesia Use |  |  |  | 0.69 |  |
| 23 | Bladder Dysfunction |  |  |  | 0.72 |  |
|  | **5. Specific Motor Symptoms** |  |  |  |  |  |
| 12 | Dystonia |  |  |  |  | 0.77 |
| 13 | Speech/aphonia |  |  |  |  | 0.60 |
| 14 | Swallowing problems |  |  |  |  | 0.43 |
| 17 | Blurred/Abnormal Vision |  |  |  |  |  |
| 11 | Tremor |  |  |  |  |  |
| 22 | Non-Epileptic Seizures |  |  |  |  |  |
| 24 | GI Dysfunction |  |  |  |  |  |

### Concurrent Validity

Data from the FiND arm were used to establish concurrent validity by means of Pearson’s correlation coefficient. Several items did not have concurrent scales conducted on the same patients. Thus, a limited number of items were available to be assessed against similar scales.

As expected, most items assessed for concurrent validity with their corresponding scales displayed strong correlation (0.62-0.91), especially Items 1 ‘Depressed Mood’ and 2 ‘Anxiety’. There was significant moderate correlation at admission and strong correlation at discharge with Items 6-8 and the WSAS. Item 9 ‘Problems with Relationships’ was weakly correlated with the WSAS. Some correlations were not significant, likely due to the smaller sample size available for the PHQ-9, GAD-7 and WSAS.

**Table 6.**
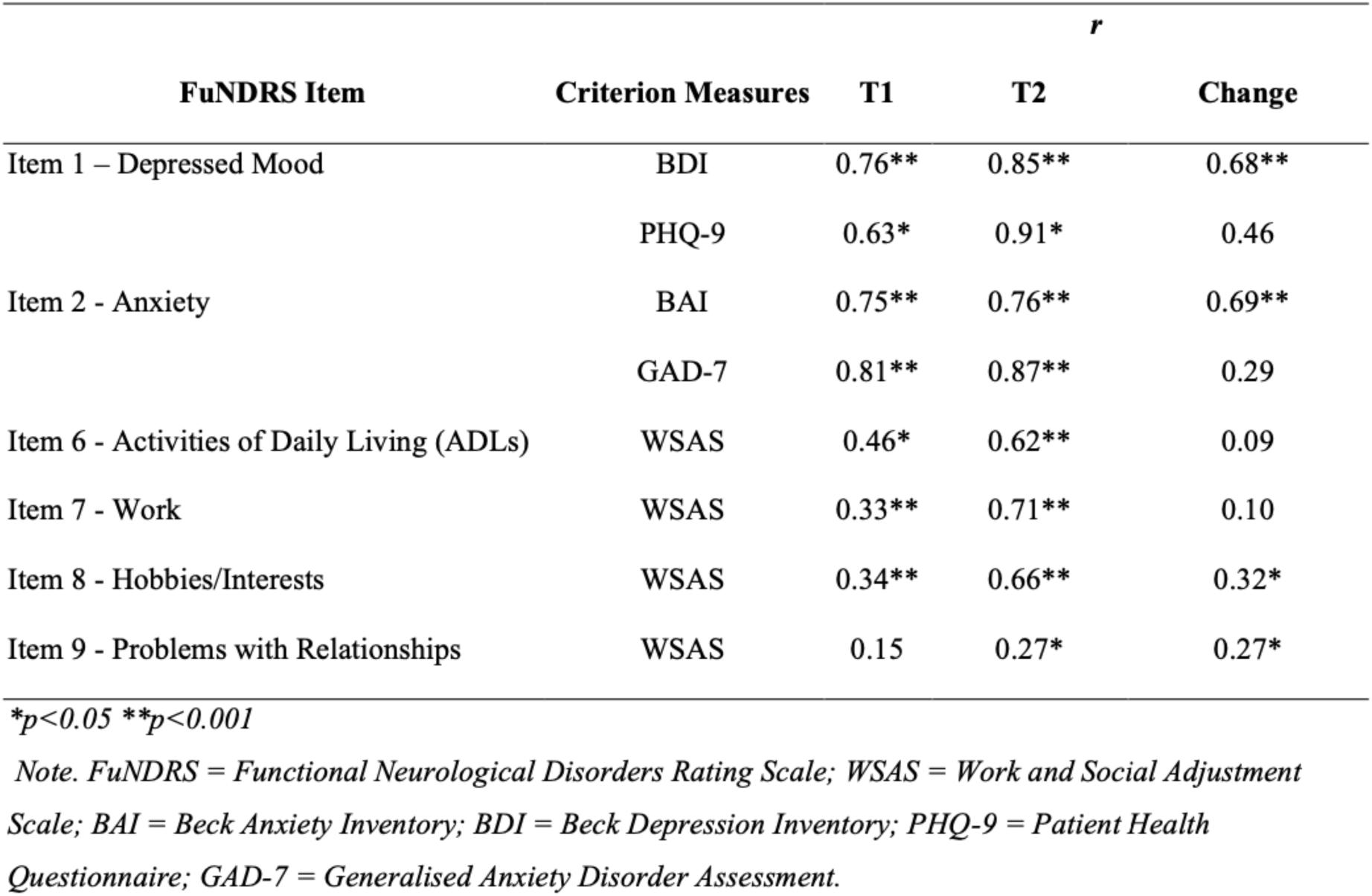
Concurrent Validity. Correlation between FuNDRS Items and criterion measures at T1 (admission), T2 (discharge) and change in score from T1 to T2. (FiND arm)

| FuNDRS Item | Criterion Measures | *r* |  |  |
| --- | --- | --- | --- | --- |
|  |  | T1 | T2 | Change |
| Item 1 – Depressed Mood | BDI | 0.76** | 0.85** | 0.68** |
|  | PHQ-9 | 0.63* | 0.91* | 0.46 |
| Item 2 - Anxiety | BAI | 0.75** | 0.76** | 0.69** |
|  | GAD-7 | 0.81** | 0.87** | 0.29 |
| Item 6 - Activities of Daily Living (ADLs) | WSAS | 0.46* | 0.62** | 0.09 |
| Item 7 - Work | WSAS | 0.33** | 0.71** | 0.10 |
| Item 8 - Hobbies/Interests | WSAS | 0.34** | 0.66** | 0.32* |
| Item 9 - Problems with Relationships | WSAS | 0.15 | 0.27* | 0.27* |

\* $p < 0.05$ \*\* $p < 0.001$
*Note.* FuNDRS = Functional Neurological Disorders Rating Scale; WSAS = Work and Social Adjustment Scale; BAI = Beck Anxiety Inventory; BDI = Beck Depression Inventory; PHQ-9 = Patient Health Questionnaire; GAD-7 = Generalised Anxiety Disorder Assessment.

### Sensitivity to Change

Paired data for admission (T1) and discharge (T2) collected from a total of 98 participants was used to assess the responsivity of the FuNDRS. As expected, patients scored lower at discharge (M = 28.72 SD = 12.19) than admission, ( M = 41.13 SD 11.07), reflecting an average difference of -12.41 (SE = 0.83) BCa 95% CI [10.77,14.05], which was significant, t(97) = 14.99, p<0.001, and resulted in a very large effect size of d = 8.19 (Cohen, 1992). Separate data for each arm are also available below.

The HoNOS (n=59) also yielded a significant change from admission (M=16.18 SD 4.8) to discharge (M=10.42 SD 5.57), reflecting an average difference of -5.77 (SE = 0.5) BCa 95% CI [4.78,6.76], which was significant, t (70) = 11.62, p<0.001, d = 4.18.

All items and domains had at least medium effect sizes, although items 23 and 24 were not statistically significantly, likely as patients scored low on these items at admission. Large effect sizes are present for 9 of 25 items (Item 1 ‘Depressed Mood’, Item 2 ‘Anxiety’, Item 5 ‘Self-Directed Injury’, Item 7 ‘Work’, Item 10 ‘Weakness’, Item 13 ‘Speech/Aphonia’, Item 15 ‘Gait/Mobility’, Item 21 ‘Analgesia’, and Item 23 ‘Bladder’). Complete data for sensitivity to change is available in Table 7.

The MDC95 was calculated by the following formula: Standard Error of Measurement X 1.96 X √2 (Ries et al., 2009), indicating a minimum change of 2.3 points correlates with a noticeable change in functional ability on the FuNDRS with 95% confidence.

**Table 7.** Sensitivity to Change of FuNDRS items at admission (T1), discharge (T2) and change in score from Tl to T2. Combined FiND and Lishman arms (n = 98).

| FuNDRS Item |  | T1<br>M | T1<br>SD | T2<br>M | T2<br>SD | Change<br>M | Change<br>SE | p* | ES |
| --- | --- | --- | --- | --- | --- | --- | --- | --- | --- |
| 1 | Depressed Mood | 2.66 | 1.07 | 1.71 | 1.32 | 0.95 | 0.12 | <0.001 | **1.15** |
| 2 | Anxiety | 2.93 | 0.91 | 2.12 | 1.16 | 0.81 | 0.10 | <0.001 | **0.95** |
| 3 | Depersonalisation/Derealisation | 1.11 | 1.36 | 0.79 | 1.09 | 0.33 | 0.06 | <0.001 | 0.64 |
| 4 | Cognitive | 1.83 | 1.20 | 1.23 | 1.10 | 0.59 | 0.07 | <0.001 | 0.73 |
| 5 | Self-directed injury | 0.57 | 1.04 | 0.38 | 0.78 | 0.19 | 0.08 | 0.02 | **0.80** |
| 6 | Problems with ADL | 2.36 | 1.00 | 1.46 | 0.95 | 0.90 | 0.08 | <0.001 | 0.78 |
| 7 | Work | 3.50 | 0.89 | 2.92 | 1.26 | 0.58 | 0.09 | <0.001 | **0.88** |
| 8 | Hobbies/interests | 3.02 | 0.71 | 2.14 | 0.97 | 0.88 | 0.08 | <0.001 | 0.75 |
| 9 | Problems with relationships | 1.86 | 1.24 | 1.53 | 1.19 | 0.33 | 0.07 | <0.001 | 0.65 |
| 10 | Weakness | 2.18 | 1.27 | 1.28 | 1.06 | 0.91 | 0.10 | <0.001 | **0.95** |
| 11 | Tremor | 0.92 | 1.31 | 0.60 | 0.96 | 0.32 | 0.07 | <0.001 | 0.70 |
| 12 | Dystonia | 0.77 | 1.35 | 0.45 | 0.92 | 0.32 | 0.07 | <0.001 | 0.73 |
| 13 | Speech/aphonia | 0.98 | 1.31 | 0.56 | 0.91 | 0.42 | 0.09 | <0.001 | **0.87** |
| 14 | Swallowing problems | 0.17 | 0.63 | 0.05 | 0.26 | 0.12 | 0.05 | 0.02 | 0.52 |
| 15 | Gait/mobility | 2.63 | 1.13 | 1.63 | 1.13 | 1.00 | 0.10 | <0.001 | **0.97** |
| 16 | Loss of sensation/numbness | 1.35 | 1.47 | 1.02 | 1.24 | 0.33 | 0.07 | <0.001 | 0.73 |
| 17 | Vision | 0.32 | 0.73 | 0.16 | 0.47 | 0.15 | 0.04 | 0.00 | 0.44 |
| 18 | Sensitivity to light/sound | 1.10 | 1.31 | 0.72 | 0.88 | 0.38 | 0.07 | <0.001 | 0.70 |
| 19 | Dizziness | 0.81 | 1.14 | 0.52 | 0.80 | 0.29 | 0.07 | <0.001 | 0.67 |
| 20 | Pain | 2.93 | 1.08 | 2.13 | 1.04 | 0.80 | 0.08 | <0.001 | **0.82** |
| 21 | Analgesia | 1.58 | 1.30 | 1.28 | 1.13 | 0.29 | 0.08 | 0.00 | 0.79 |
| 22 | Non-Epileptic Seizures | 1.55 | 1.74 | 0.92 | 1.28 | 0.63 | 0.10 | <0.001 | **1.01** |
| 23 | Bladder | 0.54 | 1.24 | 0.51 | 1.18 | 0.03 | 0.04 | 0.47 | 0.42 |
| 24 | GI | 0.94 | 1.25 | 0.87 | 1.22 | 0.07 | 0.06 | 0.21 | 0.56 |
| 25 | Daytime fatigue | 2.63 | 0.93 | 1.80 | 0.93 | 0.84 | 0.08 | <0.001 | 0.78 |
| FuNDRS Domain |  | T1<br>M | T1<br>SD | T2<br>M | T2<br>SD | Change<br>M | Change<br>SE | p* | ES |
| 1 | Mental State | 1.82 | 0.59 | 1.28 | 0.79 | 0.54 | 0.06 | <0.001 | 0.60 |
| 2 | Impact | 2.68 | 0.69 | 2.02 | 0.85 | 0.67 | 0.05 | <0.001 | 0.53 |
| 3 | Motor | 1.29 | 0.56 | 0.79 | 0.47 | 0.50 | 0.05 | <0.001 | 0.51 |
| 4 | Sensory | 1.34 | 0.59 | 0.97 | 0.52 | 0.37 | 0.04 | <0.001 | 0.35 |
| 5 | Associated | 1.42 | 0.79 | 1.02 | 0.72 | 0.39 | 0.04 | <0.001 | 0.38 |

*ES, Cohen's d ( $d < 0.20$ = small effect; $d = 0.20-0.50$ = medium effect and $d > 0.80$ = large effect.)*
*Large effect sizes indicated in bold. M = mean. SD = Standard Deviation. SE = Standard Error.*
*\*Paired sample t-tests*

### Inter-Rater Reliability

An inter-rater reliability analysis was conducted on 18 patients in the Lishman unit at admission and discharge using the FuNDRS, CGI-S and CGI-I scales. Weighted kappa (κw) with quadratic weights (Cicchetti & Allison, 1971) was run to determine if there was agreement. There was statistically significant agreement found between the two raters across all scales. Substantial agreement occurred with the FuNDRS Change, CGI-S Change and CGI-I scales. ‘Fair to moderate’ agreement was present for scales at admission and discharge for the FuNDRS and CGI-S respectively. This disparity may be reflected by the FuNDRS being a tool for outcome measurement and thus change rather than a cross-sectional measurement alone.

**Table 8.** Inter-rater reliability analysis using weighted kappa with quadratic weights. N = 18.

|  | **Weighted**<br>**Kappa** | **Lower Bound**<br>**(95% CI)** | **Upper Bound**<br>**(95% CI)** | **SE** | **p** |
| --- | --- | --- | --- | --- | --- |
| FuNDRS T1 | 0.39 | 0.13 | 0.65 | 0.13 | 0.00 |
| FuNDRS T2 | 0.33 | 0.10 | 0.56 | 0.12 | 0.04 |
| FuNDRS Change | 0.78 | 0.62 | 0.94 | 0.08 | 0.00 |
| CGI-S T1 | 0.50 | 0.14 | 0.86 | 0.19 | 0.04 |
| CGI-S T2 | 0.57 | 0.26 | 0.88 | 0.16 | 0.02 |
| CGI-S Change | 0.76 | 0.56 | 0.95 | 0.10 | 0.00 |
| CGI-I | 0.64 | 0.35 | 0.93 | 0.15 | 0.01 |

*Weighted Kappa interpreted according to Landis & Koch, (1977). (<0.2 = Slight; 0.2-0.4 = Fair; 0.4-0.6 = Moderate; 0.6-0.8 = Substantial; >0.8 = Almost Perfect).*
*Note. FuNDRS = Functional Neurological Disorders Rating Scale; HoNOS = Health of Nation Outcomes Scale; CGI = Clinical Global Impression (Scale); CGI-S = Clinical Global Impression – Severity; CGI-I = Clinical Global Impression – Global Improvement; FiND = Functional Integrated Neurological Disorders (outpatient day programme).*

Agreement was ‘almost perfect’ (>0.8) for 2 items at admission and discharge (Item 21 ‘Analgesia Use’ and Item 23 ‘Bladder Symptoms’) and for Item 22 ‘Non-Epileptic Seizures’ and Item 5 ‘Self-Directed Injury’ at admission and discharge respectively. This likely reflects levels of details of documentation in admission and discharge summaries. In total, 7 items at admission and 6 items at discharge had an agreement rated as ‘substantial’ or higher (>0.6). Additionally, there was a lack of significant agreement for several items as seen in Table 9.

**Table 9.** Inter-rater reliability analysis of FuNDRS items at admission (T1) and discharge (T2) using weighted kappa with quadratic weights. N = 18.

| Domain | FuNDRS Item | T1 (Kw) | T1 (CI 95%) | T1 (p) | T2 (Kw) | T2 (CI 95%) | T2 (p) |
| --- | --- | --- | --- | --- | --- | --- | --- |
| I. Mental State | 1 Depressed Mood | 0.659 | [0.322-0.996] | 0.006 | 0.447 | [0.022-0.872] | 0.059 |
|  | 2 Anxiety | 0.4 | [0.193-0.607] | 0.017 | 0.15 | [-0.093-0.392] | 0.359 |
|  | 3 Depersonalisation/Derealisation | 0.699 | [0.407-0.990] | 0.004 | 0.705 | [0.475-0.934] | 0.001 |
|  | 4 Cognitive | 0.381 | [0.008-0.754] | 0.108 | 0.524 | [0.212-0.836] | 0.035 |
|  | 5 Self-directed injury | 0.338 | [-0.068-0.745] | 0.156 | 0.818 | [0.628-1.009] | <0.001 |
| II. Impact | 6 Problems with ADL | 0.571 | [0.278-0.865] | 0.015 | 0.407 | [0.143-0.671] | 0.055 |
|  | 7 Work | 0.294 | [-0.152-0.740] | 0.067 | 0.06 | [-0.259-0.380] | 0.729 |
|  | 8 Hobbies/interests | 0.431 | [0.143-0.718] | 0.074 | 0.111 | [-0.276-0.498] | 0.579 |
|  | 9 Problems with relationships | 0.4 | [0.139-0.661] | 0.05 | 0.439 | [0.184-0.693] | 0.043 |
| III. Motor | 10 Weakness | 0.442 | [-0.049-0.934] | 0.061 | 0.231 | [-0.195-0.658] | 0.314 |
|  | 11 Tremor | 0.668 | [0.310-1.025] | 0.003 | 0.455 | [-0.05-0.914] | 0.033 |
|  | 12 Dystonia | 0.44 | [0.052-0.828] | 0.017 | 0.512 | [0.096-0.928] | 0.019 |
|  | 13 Speech/aphonia | 0.017 | [-0.266-0.3] | 0.903 | 0.556 | [0.194-0.917] | 0.02 |
|  | 14 Swallowing problems | 0.68 | [0.296-1.064] | 0.005 | 0.657 | [0.390-0.924] | 0.002 |
|  | 15 Gait/mobility | 0.533 | [0.112-0.954] | 0.014 | 0.156 | [-0.199-0.512] | 0.417 |
| IV. Sensory | 16 Loss of sensation/numbness | 0.388 | [0.1-0.676] | 0.015 | 0.482 | [0.103-0.862] | 0.006 |
|  | 17 Vision | 0.467 | [0.048-0.886] | 0.048 | 0.17 | [-0.279-0.618] | 0.491 |
|  | 18 Sensitivity to light/sound | 0.29 | [0.01-0.580] | 0.046 | 0.091 | [-0.078-0.26] | 0.964 |
|  | 19 Dizziness | 0.253 | [-0.017-0.523] | 0.033 | -0.075 | [-0.220-0.07] | 0.403 |
|  | 20 Pain | 0.34 | [-0.65-0.745] | 0.15 | 0.051 | [-0.334-0.436] | 0.809 |
|  | 21 Analgesia | 0.865 | [0.793-0.936] | <0.001 | 0.815 | [0.69-0.939] | <0.001 |
| V. Associated | 22 Non-Epileptic Seizures | 0.938 | [0.888-0.988] | <0.001 | 0.688 | [0.390-0.986] | 0.003 |
|  | 23 Bladder | 0.954 | [0.892-1.016] | <0.001 | 0.862 | [0.669-1.055] | <0.001 |
|  | 24 GI | 0.137 | [-0.177-0.451] | 0.415 | 0.055 | [-0.144-0.253] | 0.566 |
|  | 25 Daytime fatigue | 0.103 | [-0.085-0.290] | 0.332 | -0.054 | [-0.365-0.257] | 0.721 |

Note. Kw = weighted kappa interpreted as <0 = disagreement; 0-0.2 = poor; 0.21-0.40 = fair; 0.41-0.60 = moderate; 0.61-0.80 = good; 0.81-1.0 = very good).

Additionally, a two-way mixed intra-class coefficient analysis based on consistency was conducted to further assess conformity among the two raters (Table 9). There was statistically significant ‘fair to good ‘agreement found between the two raters across all scales according to Chichetti (1994).

**Table 10.**
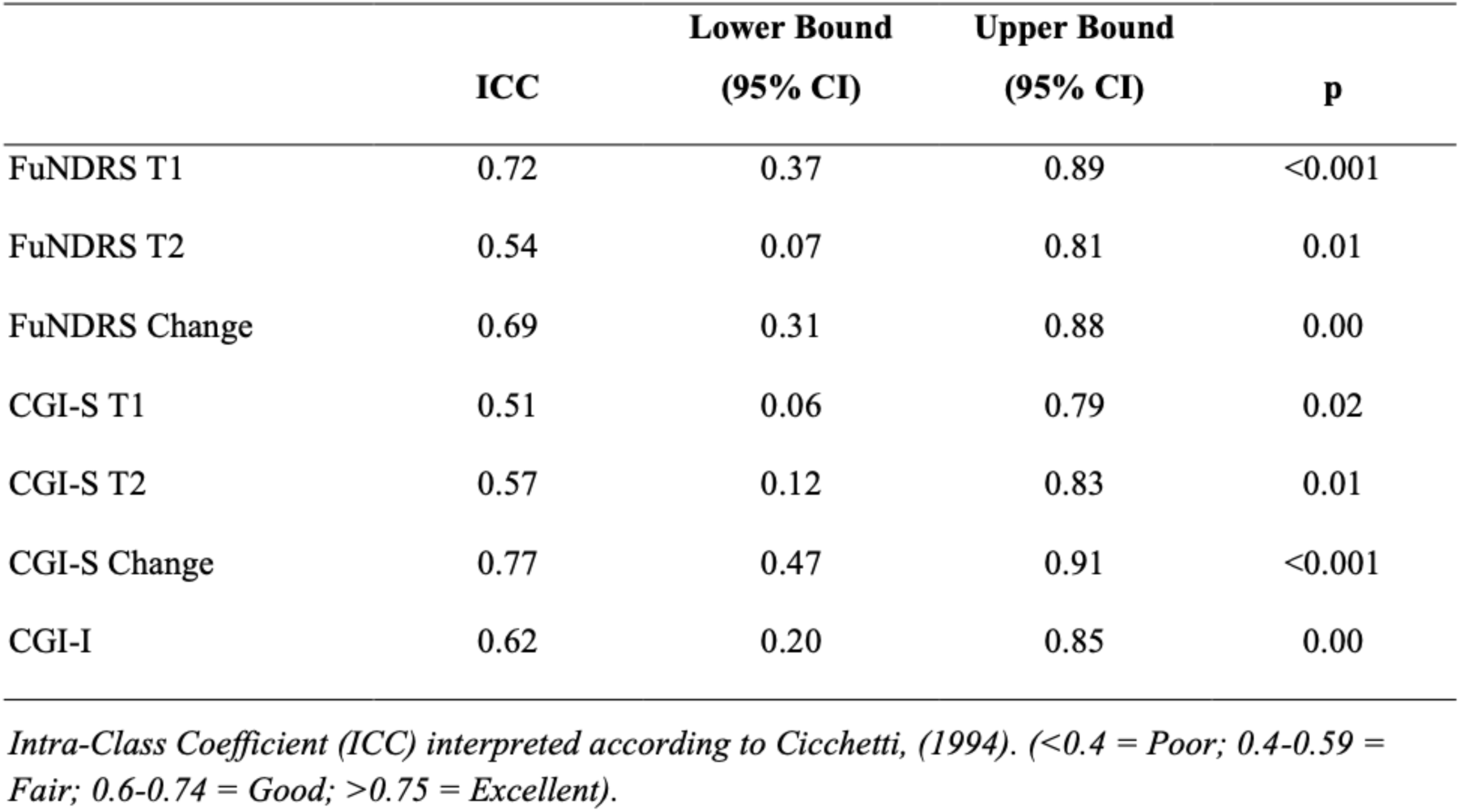
Inter-rater reliability analysis using intra-class coefficients with a two-way mixed model. N = 18.

|  | ICC | Lower Bound<br>(95% CI) | Upper Bound<br>(95% CI) | p |
| --- | --- | --- | --- | --- |
| FuNDRS T1 | 0.72 | 0.37 | 0.89 | <0.001 |
| FuNDRS T2 | 0.54 | 0.07 | 0.81 | 0.01 |
| FuNDRS Change | 0.69 | 0.31 | 0.88 | 0.00 |
| CGI-S T1 | 0.51 | 0.06 | 0.79 | 0.02 |
| CGI-S T2 | 0.57 | 0.12 | 0.83 | 0.01 |
| CGI-S Change | 0.77 | 0.47 | 0.91 | <0.001 |
| CGI-I | 0.62 | 0.20 | 0.85 | 0.00 |

*Intra-Class Coefficient (ICC) interpreted according to Cicchetti, (1994). (<0.4 = Poor; 0.4-0.59 = Fair; 0.6-0.74 = Good; >0.75 = Excellent).*

### Predictive Validity

Linearity was established by visual inspection of a scatterplot. There was independence of residuals, as assessed by a Durbin-Watson statistic of 1.609. There was homoscedasticity, as assessed by visual inspection of a plot of standardized residuals versus standardized predicted values. Residuals were normally distributed as assessed by visual inspection of a normal probability plot. A linear regression established that average FuNDRS score at admission statistically significantly predicted score at discharge, F (1, 96) = 127.84, p < .001. The regression equation was: FuNDRS score at discharge = -5.506 + 0.832 x (FuNDRS score at admission). Data for intervals between ‘10 and 70’ for FuNDRS scores at admission are presented in Table 11. All except interval ‘10’ was statistically significant.

**Table 11.** Predictive Validity of FuNDRS score at discharge using score at admission (linear regression analysis)

| **FuNDRS Score at Admission** | **Predicted FuNDRS Score at Discharge** | **SE** | **Lower Bound (95% CI)** | **Upper Bound (95% CI)** | **P** |
| --- | --- | --- | --- | --- | --- |
| 70.00 | 52.75 | 2.27 | 48.23 | 57.26 | <0.001 |
| 60.00 | 44.43 | 1.61 | 41.23 | 47.62 | <0.001 |
| 50.00 | 36.10 | 1.04 | 34.04 | 38.17 | <0.001 |
| 40.00 | 27.78 | 0.82 | 26.17 | 29.40 | <0.001 |
| 30.00 | 19.46 | 1.15 | 17.17 | 21.75 | <0.001 |
| 20.00 | 11.14 | 1.75 | 7.66 | 14.62 | <0.001 |
| 10.00 | 2.82 | 2.43 | -2.01 | 7.64 | 0.25 |

*SE = Standard Error*

## Discussion

### Overview

To our knowledge, this is the first reported pilot psychometric study for an FND outcome measurement tool and builds upon the work of a previous preliminary local validation of the FuNDRS completed by Lucy Miller. The aim was to provide initial validation for the FuNDRS across two arms using two FND populations, the Lishman Unit for inpatients and the FiND programme arm for day hospital patients. Validation was achieved through testing the FuNDRS in its following properties: convergent, discriminant, concurrent and known groups validity. Additionally, we provided data assessing the FuNDRS sensitivity to change and a preliminary factor analysis using principal components. Finally inter-rater reliability analysis was also performed on a subgroup of the inpatient arm.

### Discussion of Results

The internal consistency of the FuNDRS was good and comparable to other multidimensional scales such as the MDS-UPDRS (Goetz et al., 2008; Gallagher et al., 2012), the YBOCS (Lopez-Pina, 2015) and the UHDRS (Youssov et al., 2013). The FuNDRS performed well against the HoNOS, CGI-S and CGI-I scales, demonstrating good convergent validity. This aligns favourably with previous work by Berk and colleagues, (2008) assessing correlation between the HoNOS and CGI. Similar values are reported with the MDS-UPDRS (Goetz et al., 2008; Gallagher et al., 2012), the YBOCS (Goodman et al., 1989), and YBOCS-II (Wu et al., 2016) with their respective convergent measures. As the FuNDRS correlates strongly with the HoNOS, CGI-S and CGI-I scales, it is possible to make predictions for FuNDRS scores at discharge using linear regression, thereby demonstrating predictive validity. The range in FuNDRS scores was 53 (16 to 69) therefore appropriate intervals were presented.

Predicting FuNDRS scores at discharge may serve as a reassuring and useful tool to patients admitted to a specialised treatment programme. In turn, it can provide clinicians and MDT members confidence in the treatments they are delivering, whilst balancing reasonable and pragmatic expectations of their patients and carers at the outset.

The novel tool also showed ability to differentiate between patient groups by way of known groups validity. Furthermore, the FuNDRS displayed a sensitivity to change in a similar fashion to the YBOCS (Goodman et al., 1989) All patients except one improved across both arms and the FuNDRS was able to detect these changes reliably – arguably the most important feature supporting the validity of an outcome measurement tool (Berk et al., 2008).

Despite limited data, some FuNDRS items appear to be concurrently valid with similar scales such as items 1 ‘Depressed Mood’ and 2 ‘Anxiety’ with the BDI and PHQ-9, and BAI and GAD-7 respectively. Data was limited due to a lower sample size for PHQ-9 and GAD-7 scales, as they were introduced later in the FiND arm. Discriminant validity did not provide reassuring results, likely due to a small sample size, heterogeneity of the sample and the large overlap that exists between anxiety and depression. In the Lishman arm, the CGI correlated most with domains 1 and 2, reflecting a global clinical component embedded in the FuNDRS. In the FiND arm, there was a strong positive correlation with the HoNOS. This may suggest the HoNOS converges with the day hospital spectrum of FND whereas the CGI with the inpatient spectrum. However, the HoNOS was not conducted in the Lishman Unit, and thus more data is required before speculation regarding convergences of different patient groups can be made. Our study found no statistically significant difference between mean change in scores on the FuNDRS in the FiND arm compared with the Lishman Unit arm.

### Inter-rater Reliability

Inter-rater reliability analysis showed substantial agreement between changes in scores on the FuNDRS, CGI-S and CGI-I. There was fair to moderate agreement for the other ratings, and the FuNDRS and CGI showed a correlation despite lower agreement. This reflects FuNDRS being a tool for outcome measurement, and thus change, rather than a cross-sectional measure alone. A few limitations reduce the generalisability of this data. Firstly, a sample size of 18 can be considered low however, each patient had ratings at admission and discharge, raising the total level of observations to 36. Also, inter-rater reliability has been demonstrated with lower samples. The Parkinson’s Impulse-Control Scale (PICS) had a sample of 12 and adequately demonstrated substantial agreement (Okai et al., 2016).

Secondly, ratings were retrospectively performed from admission and discharge summaries. The second rater (PS) had no contact with the patients and independently completed the ratings blind. However, the first rater (NM) had close contact with all patients and was the responsible clinician during the inpatient admissions to the Lishman Unit. This possibly reflects the variance in agreement among items as the level of detail in documentation could vary greatly. For example, self-directed injury at discharge is often clearly commented on as it is often a criterion for discharge and mental health professionals routinely perform and document risk assessments on all patients. Another example, is analgesia use which had very high agreement, likely owing to the diligent documentation of clerking and discharging doctors.

Further contemporaneous assessment in a larger sample could further elucidate and likely strengthen agreement. As many of the therapies for FND are multimodal and involve MDT input (Espay et al., 2018); it may be useful to demonstrate the inter-rater reliability amongst psychologists, physiotherapists, occupational therapists, and specialist nurses.

### Factor Analysis

The initial factor analysis provided promising results for factor loading of certain domains, particularly as 206 observations were available for analysis. Although 3 items were excluded from the final analysis, the remaining factors explained 53.4% of the variance. It is possible that further factor analysis is required to load items more precisely onto factors, but more likely as the FuNDRS represents a multidimensional scale, it will be used as another guide to inform appropriate item inclusion. This was a similar conclusion the task force on MDS UPDRS arrived at following several iterations of factor analysis to produce clinically meaningful rotations and factor loadings (Goetz et al., 2008).

### Strengths and Limitations

Given the unblinded nature of the assessments (with exception of the second rater in the inter-rater reliability ratings) and the purposeful sampling of patients with FND, caution is necessary in relation to interpreting sensitivity to change data. However, an adequate sample size across two patient populations (inpatients and day hospital patients) and extensive validation were strengths of this study.

The lower number of similar and divergent scales with their subsequent smaller sub-sample size, limited our demonstration of concurrent validity and discriminant validity. This could be further explored with similar and divergent scales. For example, item 3 ‘Depersonalisation/Derealisation’ can be correlated with the Cambridge Depersonalisation Scale (Sierra & Berrios, 2000) and item 15 ‘Gait/mobility’ can be correlated with the ‘Timed Up and Go Test’ (Podsiadlo & Richardson, 1991).

Although external validity was not explicitly captured in this study, the data was collected in pragmatic real-world settings involving many mental health professionals across disciplines. Additionally, this data is presented for both inpatients and day hospital patients, representing the full spectrum of FND and thus supporting generalisability in similar settings. Further validation in various countries and languages, likely driven by a taskforce or large working group, would be essential to demonstrate that the FuNDRS can compete as a unified scale such as the MDS-UPDRS (Goetz et al., 2008). FND represents a heterogenous disorder, and so it is not expected patients will score highly in each domain, rather different patient groups will feature in different domains. This was especially true of non-epileptic seizures as although the CGI-S scores for NES correlated well, there was no correlation between the FuNDRS and NES score. This is concerning as it can often present as the most disabling feature of FND (Jones et al., 2016). Although future revisions of the FuNDRS may improve in this area, these limitations are often noted in multidimensional scales such as the MDS UPDRS (Goetz et al. 2008) and come at the expense of attempting to capture a highly varied and heterogenous disorder.

The HoNOS was originally intended to quantify improvement in mental health domains by the Royal College of Psychiatrists’ Research Unit, scoring high in independent reliability analysis (Wing et al., 1998). It has since been critiqued for focusing solely on social aspects of outcome measurement (Speak & Muncer, 2015). Others have previously suggested sequential ratings on the HoNOS are not a good method for assessing outcome (Bebbington et al., 1999), whilst Pirkis and colleagues (2005) believe it can be appropriately used for routinely measuring outcomes. Indeed, DeMartini and colleagues (2014), demonstrated the HoNOS can successfully predict patient-rated improvement in inpatient multidisciplinary treatment of FND. Our study demonstrated the FuNDRS converges highly with the HoNOS, therefore, it is possible that the social aspects contained in domain 2 ‘Impact’ may skew overall ratings. Comparing total scores on the HoNOS with each domain on the FuNDRS, may reveal if this is the case. The CGI-I also correlated well with domain 2 ‘Impact’ on the FuNDRS, therefore it is also likely that the social impact of FND contributes highly to illness severity, making this possible limitation in the HoNOS likely irrelevant. During the study it was apparent that distinctions between categories was disparate. For example, item 22 ‘NES’ was clearly defined as: “0=none; 1 minor - occasional seizures <1 month; 2 mild - NES>=once/month; 3 moderate -NES >=once/week; 4=severe NES; daily or almost daily seizures”. In contrast to items 9 ‘Problems with Relationships’ or 24 ‘GI

Dysfunction’ (See appendix 1), where raters found it more difficult to accurately rate patients. However, as this is an initial validation of the FuNDRS, precise wording can be improved dynamically through future iterations to improve standardisation of measurement. The FuNDRS may benefit from further validation. The ‘floor and ceiling effect’ is currently unknown and is important to ascertain in questionnaires concerned with evaluating health related quality of life measures (Hays et al., 1993; Goetz et al., 2008). Many of the other cited scales in this study such as the MDS-UPDRS incorporate patient-reported measures whilst the FuNDRS remains a wholly clinician-rated instrument. It remains unclear if the subjective component, by way of a self-reported or patient-rated measure, will support or confound validity. For example, one study found close associations between subjective and objective reported tremors in FND (Kramer et al., 2019) whereas another study found patients to overestimate tremors almost 20-fold (Pareés, 2012). As such there is insufficient evidence to guide research incorporating patient-related measures in FND.

## Conclusion

This study provides evidence for initial validation of the FuNDRS as an outcome measurement tool for FND across two patient populations. The FuNDRS displayed several favourable properties such as good convergent and concurrent validity, sensitivity to change with large effect sizes, a promising factor loading and good inter-rater reliability. The FuNDRS would benefit from further extensive validation and reliability analyses before recommendation as a gold-standard tool for outcome measurement in FND. FND-COM and COMET now represent driving forces in founding an international task force or large working group to adeptly provide external validation and generalisability across countries and diverse patient groups.

## Supporting information

Functional Neurological Disorders Rating Scale (FuNDRS)

## Data Availability

The data that support the findings of this study are available on reasonable request from the corresponding author, Lucy Miller.

## Appendix

### FuNDRS

(Functional Neurological Disorders Rating Scale)

The categories are;-

Part I – Mental state/mood/beliefs about illness/variability in symptoms
Part II – Impact of illness
Part III - Evaluation of motor function
Part IV – Sensory symptoms
Part V – Other associated symptoms

### FNDRS Maximum Score for Parts 1-5 is 100 (25 items, each scored 0-4)

Each item is scored out of max 4. Rate the following over the last two weeks.

0- no symptoms/problems
0- minor
0- mild
0- moderate
0- severe

### Part I - Mental state

1. Depressed mood

0- no low mood
1- minor mood changes (subclinical)
2- mild depression
3- moderate depression
4- severe depression
2. Anxiety

0- no anxiety problems
1- minor anxiety
2- mild anxiety
3- moderate anxiety
4- severe anxiety (generalised ± panic attacks plus somatic symptoms of anxiety/panic (palpitations/shortness of breath/paraesthesia)
3. Depersonalisation/derealisation

0- no concerns
1- minor non-clinical problem only
2- mild; occasional feelings of depersonalisation/derealisation
3- moderate; frequent feelings of depersonalisation/derealisation
4- severe; constant depersonalisation/derealisation
4. Cognitive problems (attention/memory/executive)

0- no concerns
1- minor non-clinical problem only
2- mild cognitive problems
3- moderate cognitive problems; some effect on day-to-day function
4- severe cognitive problems; significant effect on day-to-day function
5. Self-directed injury*

0- no problem of this kind during the period rated
1- minor; fleeting thoughts of self-harm or suicide but little or no risk during the period
2- mild risk during period; includes more frequent thoughts or talking about self-harm or suicide
3- moderate to serious risk of deliberate self-harm; includes frequent/persistent thoughts or talking about self-harm; includes preparatory behaviours, e.g., collecting tablets, self-injurious acts
4- suicide attempt and/or serious self-injurious acts during the period rated

*Any self-injurious behaviour which is not accidental, should be rated here; passive acts of self-injurious behaviour, e.g., failing to take action to avoid a life threatening situation, are included here.

### Part II - Impact of illness

6. Problems with activities of daily living

Rate the overall level of functioning safely in activities of daily living (ADL): e.g., problems with basic activities of self-care such as eating, washing, dressing,toilet, - also complex skills such as budgeting, use of transport.

0- no problems during the period rated; good ability to function effectively in all basic activities (e.g., continent - or able to manage incontinence appropriately, able to feed self and dress) and complex skills (e.g., driving or able to make use of transport facilities, able to handle financial affairs appropriately)
1- minor problems only without significantly adverse consequences e.g., untidy, mildly disorganised, some evidence to suggest a decline from previous functional level (especially with regard to complex skills) but still able to cope effectively
2- mild self-care and basic activities adequate though prompting may be required, but difficulty with more complex skills (e.g., problems organising and making a drink/meal, problems with driving, transport or financial judgements)
3- moderate problems evident in one or more areas of basic self-care activities (e.g., needs some supervision with dressing and eating, occasional urinary incontinence or continent only if toileted), inability to perform several complex skills in safety. Consistently requires prompting to perform activities
4- severe disability or incapacity in all or nearly all areas of basic and complex skills, or lack of safety in any area (e.g., full supervision required with dressing and eating, frequent urinary ± faecal incontinence)
7. Work

0- no concerns
1- minor non-clinical problem only
2- mild; some restriction in work activities
3- moderate; restricted in almost all work-related activities but can do some work.
4- severe; unable to work
8. Hobbies/interests

0- no concerns
1- minor non-clinical problem
2- mild; some restriction in engagement with hobbies/interests
3- moderate; restricted in almost all hobbies/interests, but still occasional involvement
4- severe; unable to engage with any hobbies/interests
9. Problems with relationships
Problems associated with social relationships, identified by the patient and/or apparent to others/carers. Rate the patient’s most severe problem associated with active or passive withdrawal from, or tendency to dominate, social relationships, and/or non-supportive, destructive or self-damaging relationships.

0- no significant problems during the period
1- minor non-clinical problem
2- mild; definite problems in making, sustaining or adapting to supportive relationships (e.g., because of controlling manner, or difficult, exploitative or abusive relationships with carers), definite difficulties reported by patient/others/carers, but mild
3- moderate; persisting significant problems with relationships; moderately severe conflict or problems identified within the relationship by the patient and/or apparent to others/carers
4- severe difficulties associated with social relationships (e.g., isolation, withdrawal, conflict, abuse); major tensions and stresses (e.g., threatening breakdown of relationship)

### Part III - Motor symptoms

10. Weakness

0- no problems
1- minor non-clinical problem
2- mild weakness, still able to use affected limbs
3- moderate weakness, but can still move affected limbs
4- severe weakness/paralysis, unable to move affected limbs
11. Tremor

0- no problems
1- minor non-clinical problem
2- mild; occasional, still able to use affected limb(s)
3- moderate; frequent tremor>25% of day
4- severe; unable to use affected limb(s)
12. Dystonia

0- no problems
1- minor non-clinical problem
2- mild; still able to use affected limb(s)
3- moderate; impaired function of affected limb(s)
4- severe fixed; unable to use affected limb(s)
13. Speech/Aphonia

0- no problems
1- minor non-clinical problem
2- mild; occasional speech impairment; able to communicate without difficulty
3- moderate; frequent speech impairment present>25% of day
4- severe; constant impairment/aphonic
14. Swallowing problems

0- no problems
1- minor occasional discomfort, no impact on diet or eating behaviours
2- mild swallowing difficulty associated with some change in dietary behaviour
3- moderate difficulties associated with dietary restriction/change
4- severe complete avoidance of oral intake due to swallowing problems
15. Gait/mobility

0- no problems
1- minor non-clinical problem
2- mild; can still walk unaided
3- moderate; requires crutches/frame
4- severe; unable to walk; wheelchair user

### Part IV Sensory symptoms

16. Loss of sensation/numbness

0- no problems
1- minor non-clinical problem
2- mild; does not affect function
3- moderate; some effect on function of affected area/limb
4- severe; significant impairment of function as a result
17. Blurred/abnormal vision

0- no problems
1- minor non-clinical problem
2- mild loss of visual acuity; mild impact on function only
3- moderate visual impairment; requires assistance with some activities as a result
4- severely abnormal vision; visually impaired/blind
18. Sensitivity to light/sound

0- no problems
1- minor; non-clinical problem
2- mild; some attempts to block out light/sound
3- moderate; need to block sensory input e.g., wearing sunglasses/earplugs but still able to leave bedroom and interact with others
4- severe intolerance; avoidance e.g., unable to leave darkened room, curtains drawn etc.
19. Dizziness

0- no problems
1- minor; occasional, not clinically significant
2- mild; intermittent but severe enough to cause some behavioural change/avoidance
3- moderate with frequent avoidance of upright posture
4- severe; completely unable to maintain upright posture due to severe dizziness
20. Pain symptoms

0- no problems
1- some complaints of pain; minor non-clinical problem
2- mild pain; no effect on function
3- moderate pain; some restriction in activity as a result
4- severe pain; significant effect on activity
21. Analgesia use

0- none
1- minor non-opiate/ non-psychotropic* analgesia use
2- mild; regular low dose opiate (e.g., co-codamol) and/or psychotropic*
3- moderate; regular moderate dose (< BNF max tramadol/codeine) opiate ± psychotropic*
4- severe; regular high dose (max/supra BNF dose and/or multiple agent) opiate ± psychotropic*

*Psychotropic analgesia = duloxetine/amitriptyline/gabapentin/pregabalin

### Part V-Convulsive symptoms (must be considered to be functional in aetiology rather than organic)

22. Non-epileptic seizures

0- none
1- minor; occasional seizures <1 month
2- mild; NES>=once/month
3- moderate; NES >=once/week
4- severe NES; daily or almost daily seizures

### Part VI – associated symptoms

23. Bladder dysfunction

0- none
1- minor; non-clinical problem
2- mild; occasional mild dysfunction (incontinence/retention/frequency/dysuria)
3- moderate; frequent (but less than daily symptoms) dysfunction
4- severe; daily or constant symptoms; catheterised or self-catheterises
24. GI dysfunction (includes nausea/vomiting/ IBS symptoms/ constipation

0- none
1- non-clinical problem
2- minor; occasional complaints of GI symptoms
3- moderate; frequent complaints of symptoms; requesting or taking medical treatment (anti-emetics/laxatives etc.)
4- severe; daily or constant symptoms; multiple medical treatments; PEG fed/enema use
25. Daytime fatigue

0- none
1- minor non-clinical problem
2- mild; able to continue with activities
3- moderate; some effect on activities
4- severe; major restriction in activity level as a result; housebound or bedbound

